# Factors associated with COVID-19 susceptibility and severity in patients with multiple sclerosis: A systematic review

**DOI:** 10.1101/2021.06.11.21258765

**Authors:** Mahdi Barzegar, Sara bagherieh, Shakiba Houshi, Mozhgan Sadat Hashemi, Ghasem Pishgahi, Alireza Afshari-Safavi, Omid Mirmosayyeb, Vahid Shaygannejad, Aram Zabeti

## Abstract

**Background:** We conducted this systematic review to identify factors associated with coronavirus disease (COVID-19) susceptibility and outcomes among people with multiple sclerosis (MS).

**Methods:** Available studies from PubMed, Scopus, EMBASE, Web of Science, and gray literature including reference list and conference abstracts were searched from December 1, 2019, through April 12, 2021. We included cross-sectional, case-control, and cohort studies that reported risk factors of contracting COVID-19 or its outcome in patients with MS on univariate or multivariate regression analyses.

**Results:** Out of the initial 2719 records and 1553 conference abstracts, a total of 20 studies were included. Factors associated with COVID-19 susceptibility were reported in 11 studies and risk factors for infection outcomes were discussed in 10. History of contact with an infected is strongly suggested as a risk factor for COVID-19 susceptibility. Other factors that could be associated with contracting infection are younger age, relapsing course, and anti-CD20 agents. The evidence suggests that increasing age, greater MS severity, treatment with anti-CD20 agents, previous use of corticosteroids, and specific comorbidities (obesity and coronary artery disease) could be independently associated with worse infection outcomes. Male sex is likely to be a risk factor for more severe disease. The black or African American race was reported as a possible risk factor.

**Conclusion:** Due to a paucity of research and methodological issues, no risk factors for COVID-19 susceptibility and outcomes neither be confirmed nor excluded. Further large studies are needed to address factors associated with COVID-19 susceptibility and severity.

## 1. Introduction

The outbreak of Coronavirus Disease-2019 (COVID-19), caused by the SARS-CoV2 Virus, has led to a newly emerging pandemic. This globally-spreading virus affects people in different ways, with manifestations ranging from no symptoms to hospitalization and death due to Acute Respiratory Distress Syndrome (ARDS) [1]. More than one year after the outbreak of COVID-19, the number of reported COVID-19 cases exceeds 150 million, with more than 3.5 million deaths [2].

Multiple sclerosis (MS) is one of the most common demyelinating diseases in the central nervous system (CNS), affecting generally young female adults. Because MS patients often receive immunosuppressive agents and the disease is associated with several comorbidities [3, 4], a question was raised regarding whether people living with MS were at higher risk of COVID-19 and more likely to develop severe symptoms when infected. With the continuation of the coronavirus pandemic, an increasing number of studies have described the clinical features and outcomes of COVID-19 among MS patients [5-7]. The pooled prevalence of COVID-19 was estimated as 4% [8]. A recent systematic review has suggested a mortality rate of 3.5% among suspected/confirmed COVID-19 cases, which is slightly higher than the rate of 2.2% among the general population [9].

Current evidence suggests that aging, male sex, smoking, race/ethnicity, and underlying comorbidities such as malignancy, chronic pulmonary diseases, chronic kidney diseases, diabetes, heart diseases, hypertension, smoking, and obesity could be risk factors for severe COVID-19 [10, 11]. However, there is still no overall conclusion on the factors associated with COVID-19 in patients with MS. Such information is useful for identifying strategies to improve care for MS patients to prevent the development of severe COVID-19 infection and its associated complications. Therefore, this systematic review was conducted to present the current evidence regarding risk factors of COVID-19 susceptibility and outcome among people living with MS.

## 2. Method

### 2.1. Inclusion and exclusion criteria

Studies were included according to the following criteria: population (participants), outcomes, and study types. Population (participants): suspected or confirmed COVID-19 patients with a previous diagnosis of MS. Outcomes: risk factors for COVID-19 susceptibility and outcomes reporting on univariate or multivariate regression analyses. Study types: cross-sectional, case-control, and cohort studies. Studies with the following characteristics were excluded: (a) studies investigated specific DMT, (b) non-peer-reviewed articles, (c) non-English studies, and (d) review articles, systematic review, and qualitative studies.

### 2.2. Information source and search strategy

We comprehensive searched electronic databases including PubMed, Scopus, EMBASE, and Web of Science from December 1, 2019, to April 12, 2021. The following search words were adapted: ((coronavirus OR Wuhan coronavirus OR novel coronavirus OR coronavirus disease OR COVID-19 OR 2019 novel coronavirus infection OR 2019-nCOV OR severe acute respiratory syndrome coronavirus 2 OR SARS-CoV-2) AND (Multiple Sclerosis OR (Sclerosis, Multiple) OR (Sclerosis, Disseminated) OR Disseminated Sclerosis OR (Multiple Sclerosis, Acute Fulminating)). We also screened the reference lists of identified articles, review studies, or other relevant documents for inclusion in the study. In addition, we also searched the online library and abstracts of the following congresses: 8^th^ American and European Committee for Treatment and Research in Multiple Sclerosis (ACTRIMS-ECTRIMS 2020), 145^th^ Annual Meeting American Neurological Association, and 6^th^ Congress of the European Academy of Neurology to identify eligible studies that have not been published. We conducted this systematic review following the Preferred Reporting Items for Systematic Reviews and Meta-Analyses (PRISMA) guidelines [12].

### 2.3. Study selection

Two researchers (MB and SB) independently screened the titles and abstracts of retrieved studies to identify the eligible studies. Then, the full text of the potentially eligible studies was reviewed. Disagreement regarding the study selection was resolved by consulting with a third investigator (AAS).

### 2.4. Quality assessment

Two reviewers (OM and MB) independently evaluated the quality of the included studies using the Newcastle-Ottawa scale (NOS) quality tests [13]. Different checklists were used based on the study design. The third investigator solved any discrepancies (AAS) in quality assessment.

### 2.5. Data extraction

Two researchers (MSH and GP) independently carried out the extraction of data. The following information was extracted from each eligible publication: first author’s name, initial publication date, location of study, type of study, the total number of patients with MS, number of MS patients with confirmed/suspected COVID-19, age, sex, the severity of MS, duration of MS, course of disease (relapsing-remitting MS [RRMS], progressive), DMT classes, comorbidity (cardiovascular disease, diabetes mellitus, hypertension, chronic lung diseases, liver diseases, kidney diseases, malignancy, smoking status, obesity, and BMI score), previous corticosteroid use, vitamin status, socioeconomic status, history of contact with infected case, history of contact with a person with respiratory symptoms. Any other risk factor reported in at least one study was also extracted.

## 3. Results

### 3.1. Study selection

A total of 2719 records were initially identified according to the research strategy. After duplicate removal, 2443 retrieved studies were screened in the title and abstract. Among 126 records were reviewed in the full-text, 17 published articles met inclusion criteria. Out of 1553 conference abstracts, 3 met inclusion criteria. Finally, a total of 20 studies were included in this systematic review. The PRISMA flow chart shows the process of study selection (Figure 1). The quality assessment for each published article is presented in Table 1.

**Figure 1.**
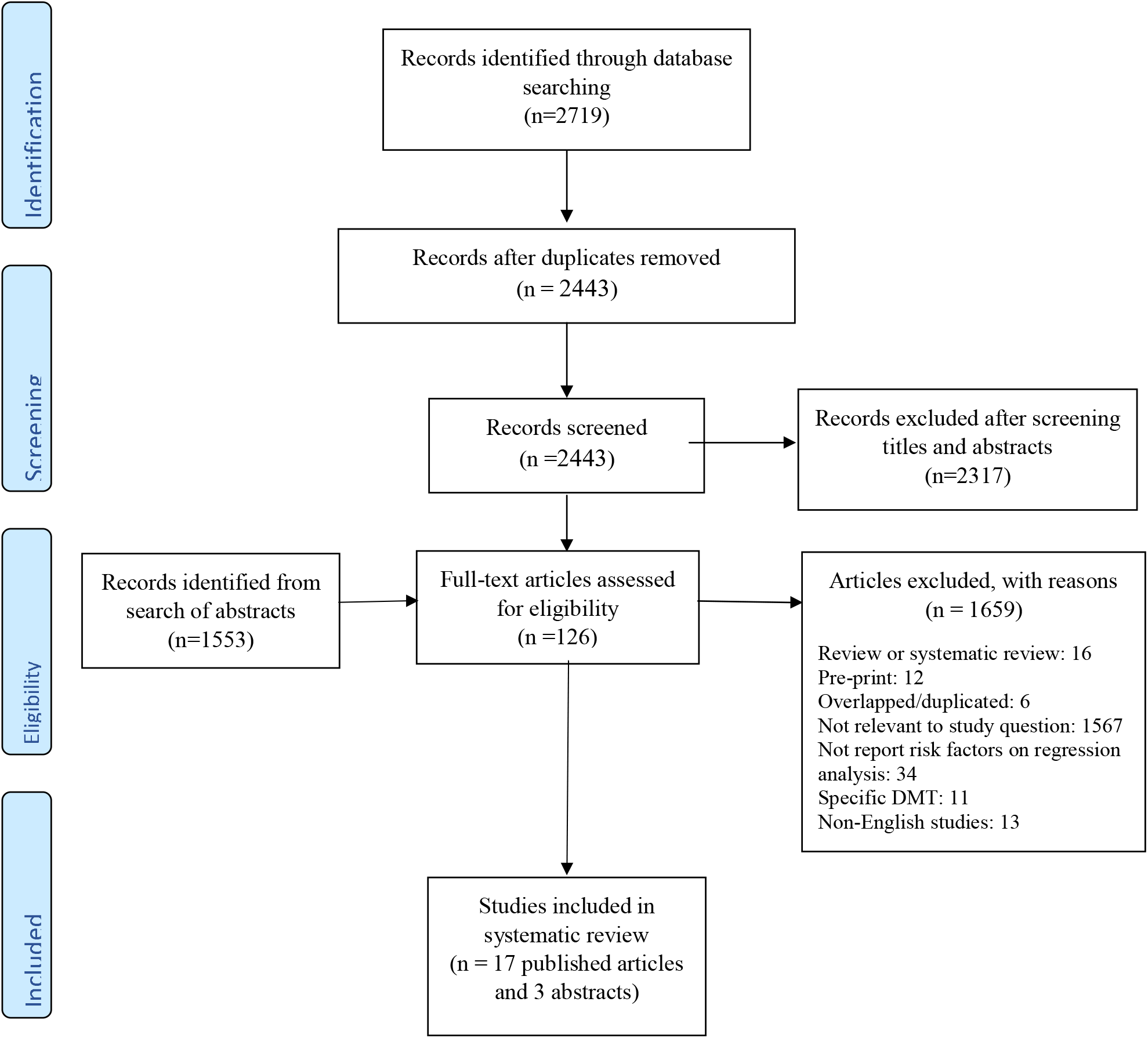
Study flowchart

**Table 1a.**
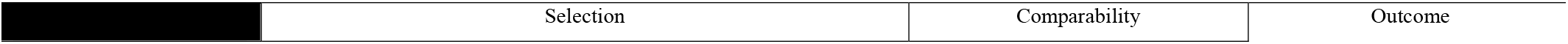
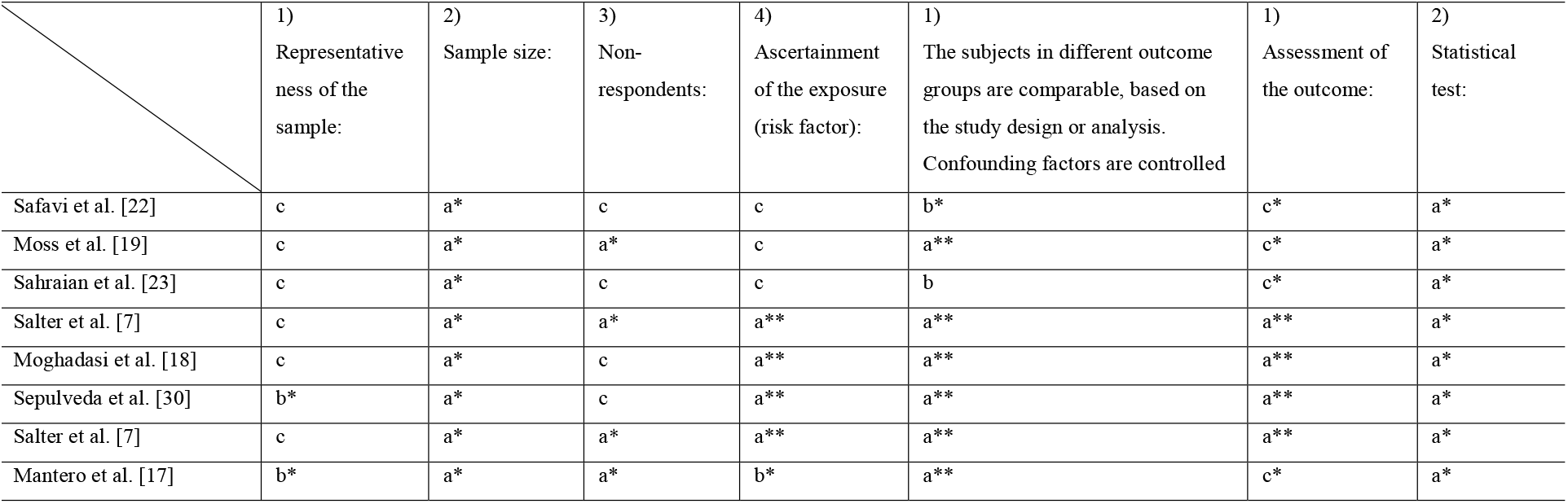
Quality assessment of cross-sectional published articles

|  | Selection | Comparability | Outcome |
| --- | --- | --- | --- |

|  | 1)<br>Representative<br>ness of the<br>sample: | 2)<br>Sample size: | 3)<br>Non-<br>respondents: | 4)<br>Ascertainment<br>of the exposure<br>(risk factor): | 1)<br>The subjects in different outcome<br>groups are comparable, based on<br>the study design or analysis.<br>Confounding factors are controlled | 1)<br>Assessment of<br>the outcome: | 2)<br>Statistical<br>test: |
| --- | --- | --- | --- | --- | --- | --- | --- |
| Safavi et al. [22] | c | a* | c | c | b* | c* | a* |
| Moss et al. [19] | c | a* | a* | c | a** | c* | a* |
| Sahraian et al. [23] | c | a* | c | c | b | c* | a* |
| Salter et al. [7] | c | a* | a* | a** | a** | a** | a* |
| Moghadasi et al. [18] | c | a* | c | a** | a** | a** | a* |
| Sepulveda et al. [30] | b* | a* | c | a** | a** | a** | a* |
| Salter et al. [7] | c | a* | a* | a** | a** | a** | a* |
| Mantero et al. [17] | b* | a* | a* | b* | a** | c* | a* |

**Table 1b.** Quality assessment of cohort published articles

|  | Selection |  |  |  | Comparability | Outcome |  |  |
| --- | --- | --- | --- | --- | --- | --- | --- | --- |
|  | 1)<br>Representative<br>ness of the<br>exposed cohort | 2)<br>Selection of<br>the non-<br>exposed<br>cohort | 3)<br>Ascertainmen<br>t of exposure | 4)<br>Demonstration that<br>outcome of interest<br>was not present at<br>start of study | 1)<br>Comparability of<br>cohorts on the basis<br>of the design or<br>analysis controlled<br>for confounders | 1)<br>Assessme<br>nt of<br>outcome | 2)<br>Was follow-up<br>long enough<br>for outcomes<br>to occur | 3)<br>Adequacy of<br>follow-up of<br>cohorts |
| Louapre et al. [5] | b* | c | a* | a* | b | a* | a* | a* |
| Evangelou et al. [15] | b* | a | c | c | b | a* | a* | b* |
| Dalla Costa et al. [14] | b* | c | c | a* | a* | a* | a* | b* |
| Alonso et al. [25] | b* | c | b* | a* | b | a* | a* | b* |
| Reder et al. [21] | b* | a* | c | a* | a* | a* | a* | b* |
| Brum et al. [26] | b* | c | a* | a* | a* | a* | a* | b* |
| Sormani et al. [6] | b* | a* | c | a* | b | a* | a* | b* |
| Zabalza et al. [24] | b* | c | c | a* | a* | a* | a* | b* |
| Levin et al. [16] | a* | c | c | a* | b | a* | a* | b* |
| Sen et al. [29] | c | a* | c | b | b | a* | a* | a* |

### 3.2. Characteristics of studies included

The characteristics of included studies are summarized in Tables 2a and 2b. Eleven studies with 46331 MS patients consisting of 1063 cases of suspected/confirmed COVID-19, investigated risk factors for COVID-19 susceptibility [14-24]. Nine studies with 4061 MS patients with suspected/confirmed COVID-19 evaluated factors associated with infection outcomes [5-7, 24-30]. One study reported both risk factors of COVID-19 and its severity [24]. Eight of the included studies were cross-sectional [7, 17-20, 22, 23, 30], 11 were cohort [5, 6, 14-16, 24-29], and one was pharmacovigilance [21]. Four studies reported data each from the USA [7, 16, 21, 27] and Iran [18, 20, 22, 23], three from Spain [24, 28, 30], two each from Italy [6, 17], and Latin America [25, 26], and one each from France [5], United Kingdom [15], and Turkey [29]. Two studies were multi-centric, one from Europe [14] and another was conducted in the USA and Spain [19].

**Table 2a.**
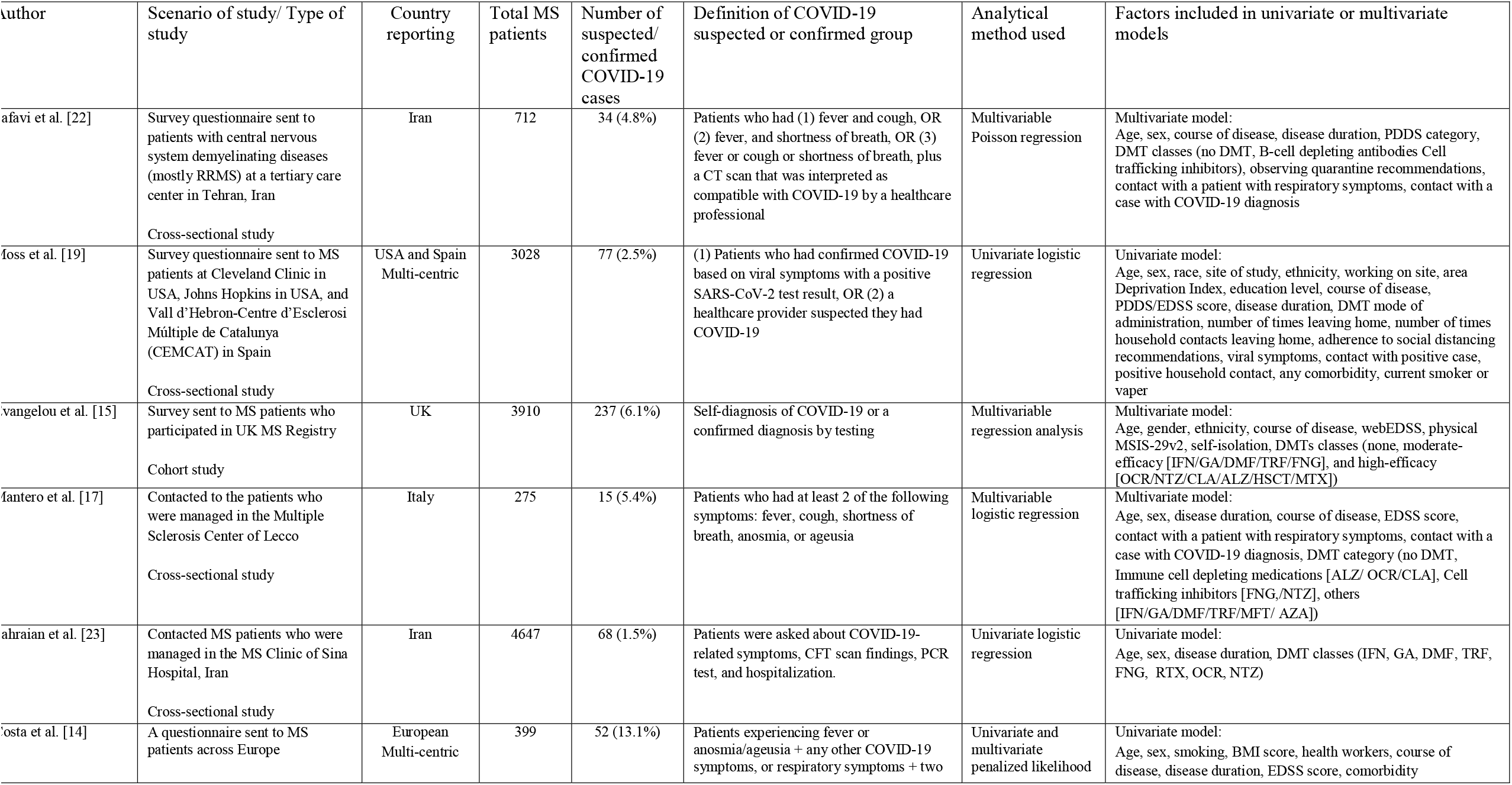
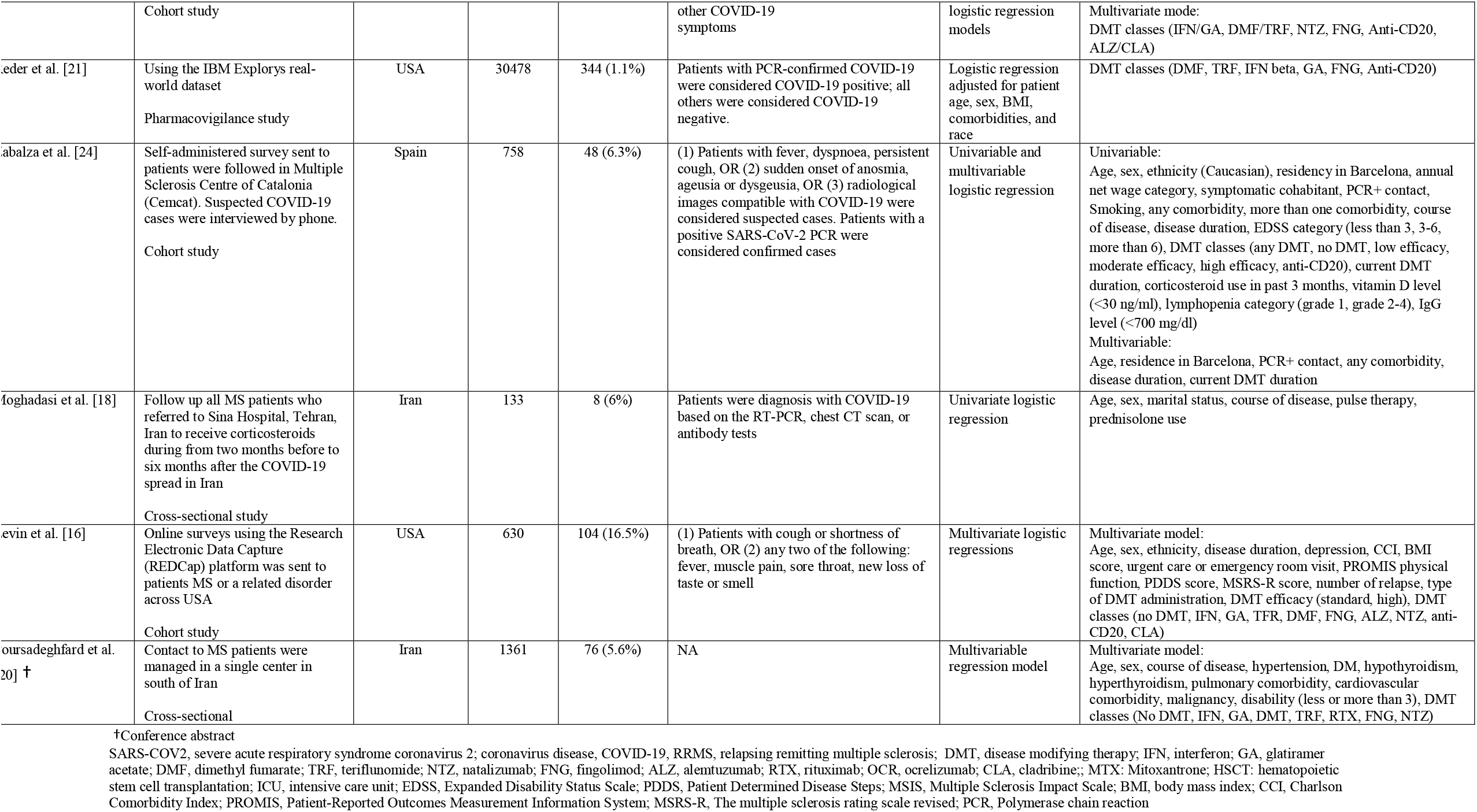
Characteristics of studies reporting risk factors of COVID-19 susceptibility

**Table 2b.**
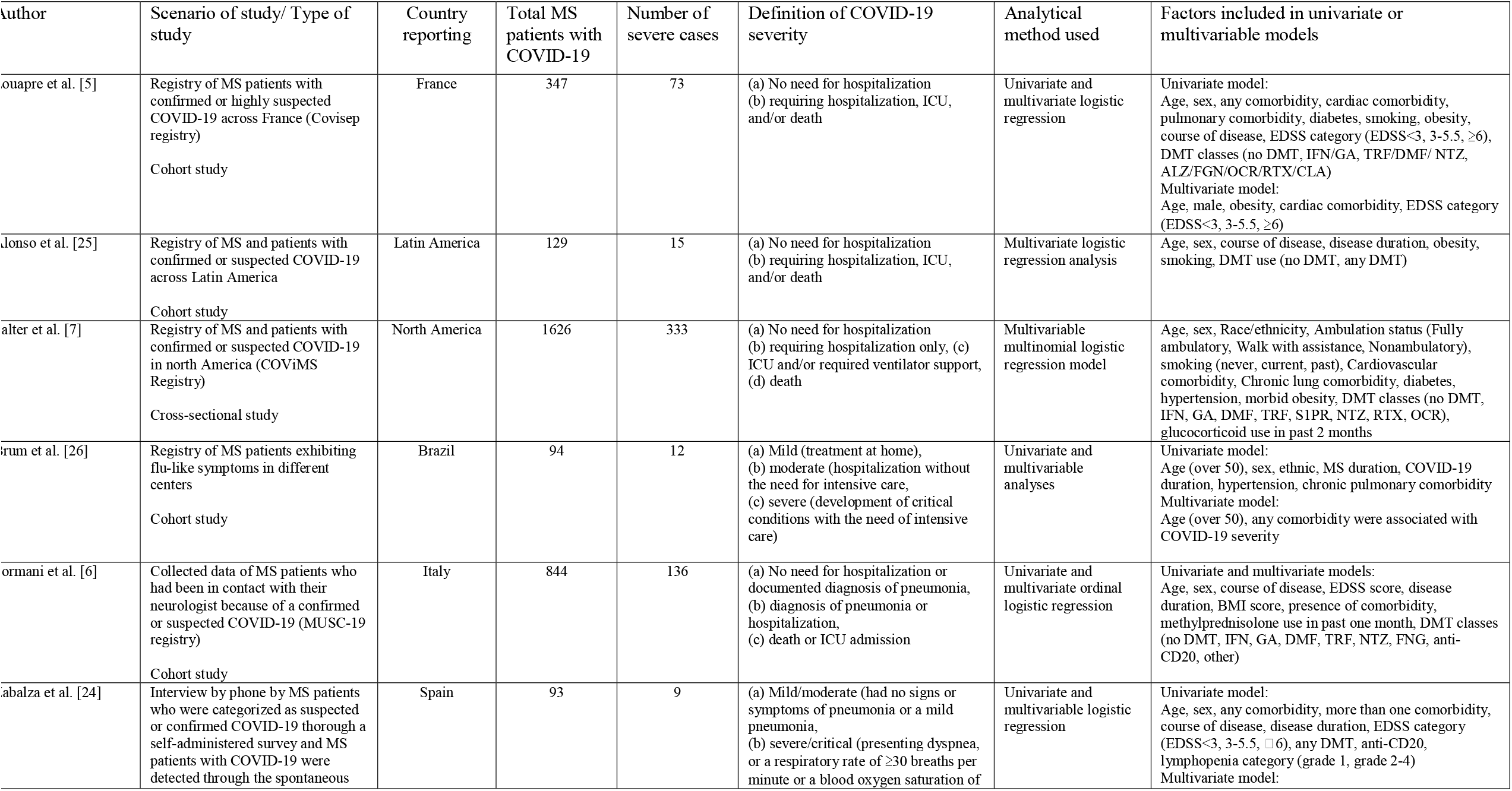
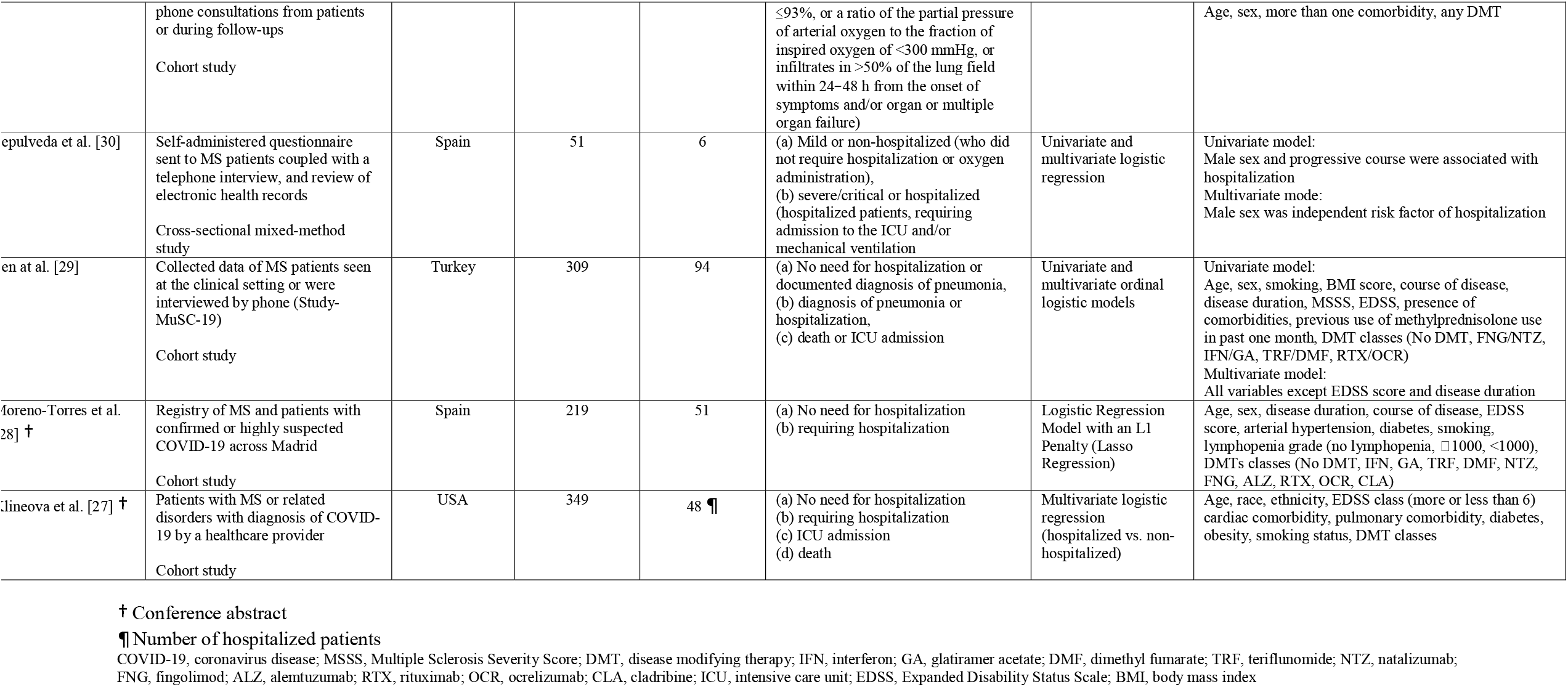
Characteristics of studies reporting risk factors of COVID-19 severity

Seven of 11 studies reported the results of multivariate analyses assessing risk factors of COVID-19 infection among MS patients. One study used a univariate model adjusting for confounders and 3 presented univariate analysis only. Nine of 10 studies considering risk factors of COVID-19 worse outcome applied multivariate analysis. Of them, 2 used the ordinal logistic regression model [6, 29] and 1 [7] conducted multinomial logistic regression. One study reported the results of only univariate analysis, with an L1 Penalty (Lasso Regression) [28].

### 3.3. Specific risk factors

We discussed each risk factor that has been considered at least in one study. For each risk factor, we initially discussed its association with susceptibility for COVID-19. Then, we reported the results of studies investigating risk factors of the COVID-19 outcome. Possible factors have been investigated in only one study discussed in the other risk factors section. One study reported risk factors of mortality [7]. In the end, we presented the conclusions emerging from the systematic review in Table 3.

**Table 3.**
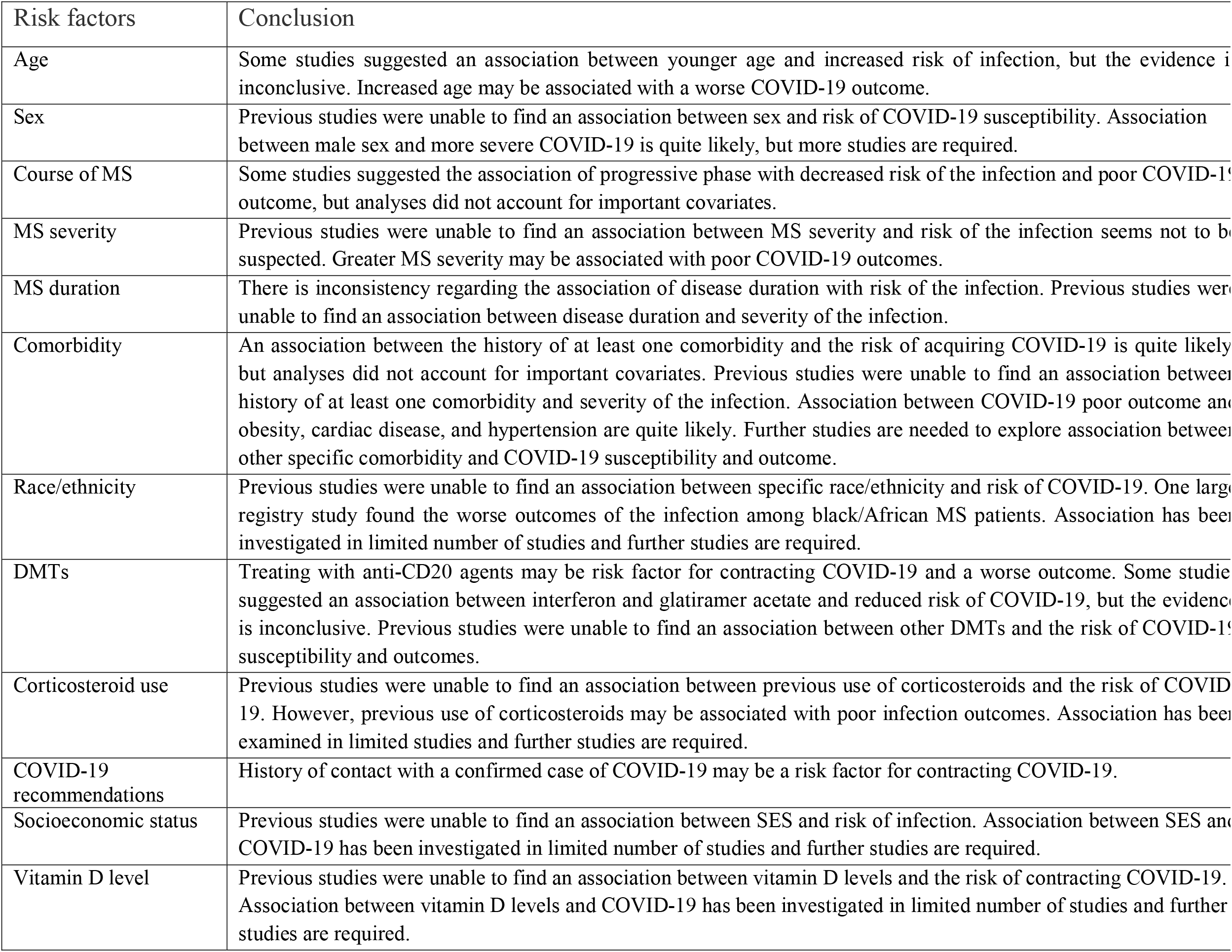
Summary of the findings

#### 3.3.1. Age

Two of the ten studies investigating the association of age with COVID-19 suggested that patients with younger age could be at increased risk of COVID-9 [OR 1.043 (1.022, 1.064) [15], OR 0.53 (0.34, 0.85) [24]], after controlling for confounders. The remaining studies have found no association between confirmed/suspected COVID-19 and age [14, 16-20, 22, 23].

Of the studies discussing age and severity of COVID-19, six suggested an independent association between greater age and elevated risk of hospitalization, with the risk per decade ranged from 6% to 171% [5-7, 24, 25]. A study showed that patients 50 years and over had 3.922 (1.383, 11.121) times the odds ratio severe COVID-19 [26]. Older age was reported as an independent risk factor for mortality [OR 3.12 (1.46-6.65)] [7]. Four studies didn’t find an association between age and severity of the disease [27-30].

#### 3.3.2. Sex

Ten studies examined sex and none provided evidence for being an independent risk factor of COVID-19 susceptibility [14-20, 22-24].

One of nine studies discussing sex and severity of COVID-19 found that male patients at higher risk of hospitalization [OR 12.56, (1.21–130.78)] [30]. The results of the French Covisep registry showed no association between male sex and hospitalization, ventilation, or death. But, they found an independent association when the severity of COVID-19 was defined as requiring supplemental oxygen therapy or higher severity [OR 2.56 (1.22, 5.35)] [5]. In the North American registry study, male patients with MS were three times more likely to die of COVID-19 compared to women [OR 3.12 (1.46-6.65)] [7]. Other six studies found no independent association between sex and severity of the infection [6, 24-26, 28, 29].

#### 3.3.3. Course of MS

Three of nine studies discussing the course of MS supported an association between the progressive phase and a decreased risk of COVID-19 [OR 0.429 (0.241 to 0.763) [15], OR 2.46 (1.72–3.20) [23], OR 0.36 (0.13–0.84) [14]]. However, only one of these studies used a multivariate model [15] and two others just conducted univariate regression analysis [23]. Other six studies have found no association between the course of the disease and confirmed/suspected COVID-19 [17-20, 22, 24].

Seven studies have investigated the association between the course of MS and the severity of COVID-19. One study found that progressive course was an independent risk factor for worse COVID-19 outcome [OR 2.36 (1.09, 5.13)] [29]. One study observed a decreased risk of hospitalization in patients with RRMS at univariate model [OR 0.24 (0.07–0.80)], but it didn’t perform a multivariate model [28]. Others found no association in the multivariate model [5, 5, 24, 25, 30].

#### 3.3.4. Severity of MS

Eight studies investigated the association of MS severity and confirmed/suspected COVID-9 [14-17, 19, 20, 22, 24]. Different methods for defining the severity of the disease have been used. One study found that MSRS-R could be an independent risk factor of suspected COVID-9 [OR 1.45 (1.11, 1.78)] [16]. This study also used the PDDS score that which wasn’t a risk factor. One study using webEDSS score and MSIS-29v2 found no association [15]. Other studies used different categorizations of EDSS and PDDS scores and found no association between severity of disease and COVID-19.

Three of nine studies discussing MS severity and outcome of COVD-19 found that ambulatory disability was independently associated with COVID-19 worse COVID-19 outcomes [5, 7, 27]. One study suggested an association between the severity of COVID-19 and MSSS ≥7 [OR 2.13 (1.07, 4.23)], but not with an EDSS score [29]. Other five studies observed no relation between MS severity and outcome of the disease [6, 6, 25, 28, 30]

#### 3.3.5. Duration of MS

Of the seven studies that included discussion of disease duration, one found that increased disease duration was independently associated with decreased risk of being suspected for the infection [OR 0.92 (0.86, 0.99)] [22]. In contrast, another study showed an association between longer disease duration and elevated risk of COVID-19 [OR per 5 years 1.41 (1.09, 1.83)] [24]. Others found no association between disease duration and suspected/confirmed COVID-19 [14, 14, 17, 19, 23].

One of seven studies discussing the association between disease duration and severity of the infection suggested that longer disease duration is an independent risk factor for hospitalization and ICU [OR 1.39 (1.14-1.69)] [25], whereas others found no relationship [6, 6, 26, 28-30].

#### 3.3.6. Comorbidity

Three studies found an association between any comorbidity and acquiring COVID-19 in univariate model [OR 1.83 (1.09, 3.07) [19], OR 2.05 (1.10, 3.80) [24], 1.03 (0.54-1.88) [14]]. However, only one applied multivariate model which result didn’t show a significant association. One study found an increased risk of COVID-19 in patients with more than one comorbidity in the univariate model, but not in the adjusted model [24]. Three studies investigated the effect of smoking on the susceptibility of MS patients to COVID-9, but none found any association [14, 19, 24]. Two studies found no specific association between BMI score and confirmed/suspected COVID-19 [14, 16]. Specific comorbidities were discussed in one conference abstract which found hyperthyroidism [OR 21.3 (2.95-55.08)] and cardiovascular disease [OR 3.69 (1.47-9.21)] as risk factors of infection [20].

None of six studies have investigated the association between the presence of at least one comorbidity and infection severity found it as an independent variable [5, 6, 24, 27, 29, 30]. One study suggested an independent association between the presence of more than one comorbidity and a higher risk of severe COVID-19 disease [OR 37.329 (2.279, 61.445)] [26], but another study didn’t find the association at the multivariate model [24]. Obesity was independently associated with worse outcomes in three studies [OR 1.69 (1.03-2.75) [7], OR 2.99 (1.03-8.70) [5], OR 2.1 (1.1, 4.1) [27]], but one didn’t find any association [25]. Two studies considered BMI scores but none found any relationship [6, 29]. One of five [5, 7, 27, 28, 30] studies suggested diabetes as an independent risk factor [OR 2.46 (1.50-4.04) [7]]. One of three [5, 7, 30] suggested an association between cardiovascular disease diseases and risk of hospitalization [OR 1.91 (1.02-3.59) [7]]. It was also found as a mortality risk factor [OR 3.15 (1.18-8.45)] [7]. No independent association between the risk of hospitalization and smoking [5, 7, 27-30] and pulmonary diseases [5, 7, 30] were observed. Hypertension was reported as an independent factor associated with mortality [OR 3.14 (1.38-7.15)] [7], but not hospitalization [7, 26, 28, 30].

#### 3.3.7. Race and ethnicity

Three studies from the USA [16], UK [15], Spain [24], and a multi-centric study on American and European MS patients [19] examined race and ethnicity. There was no association between suspected/confirmed COVID-19 and Black/African American [19], non-Hispanic Caucasian [16], Caucasian [24], and white race [15].

Three studies have discussed the association of race and ethnicity with COVID-19 clinical severity. The results North America (COViMS) registry showed a higher risk of hospitalization [OR 1.47 (0.98-2.22)] and ICU admission [OR 2.28 (1.22-4.23)] in patients with black race compared to Non-Hispanic White, but not mortality [7]. Another study from the USA [27] and a multi-centric study on Brazilian MS patients [26] found no effect for the race.

#### 3.3.8. Disease-modifying therapy

Two studies showed that patients who received anti-CD20 agents tend to be associated with an elevated risk for COVID-19 compared to other DMT groups [OR 3.6 (1.45, 8.68) [22]] and those on DMF [OR 3.25 (2.31–4.64) [21]], after adjusting for covariates. One study reported a relation between rituximab and COVID-19 diagnosis [OR 1.85 (1.37–2.33)] in the univariate model [23]. Four studies observed no association between anti-CD20 agents and COVID-19 [14, 16, 20, 24]. After adjusting for covariates, one study found a decreased risk of COVID-19 in patients on GA compared to DMF [21], and one found an inverse association between IFN and risk of contracting COVID-19 [16]. There was an increased risk of infection in patients treated with ALZ/CLA in comparison with those on INF/GA [14]. Three studies found no association between COVID-19 and IFN and GA [20, 23, 24]. Three studies have considered other specific DMT and none found an association between other DMTs and the risk of COVID-19 [14, 21, 23]. Regarding the association of no DMT with susceptibility, one of six studies [15-17, 22-24] found that high-effective DMT was associated with reduced risk of the infection compared to those on no DMT [OR 0.540 (0.311 to 0.938)] [15].

Nine studies have discussed the effect of DMTs on the outcome of COVID-19 [5-7, 24, 25, 27-30]. The results of the COViMS registry suggested that patients receiving rituximab [OR 4.56 (2.10-9.90)] and ocrelizumab [OR 1.63 (0.98-2.72)] were more likely to be hospitalized compared to those who were on no DMT, but not requiring to ICU or death [7]. In the line with this study, the Italian MUSC registry found an association between treating with anti-CD20 agents and worse outcome of COVID-19 [OR 2.37 (1.18–4.74)] [6]. Four studies found no association between anti-CD20 agents and the severity of the infection [24, 27, 29, 30]. No association between other DMTs and severity of the infection in the multivariate model were found [5-7, 24, 25, 27-30]. Three studies found worse COVID-19 outcome among those received no DMT, but this association didn’t remain significant in the adjusted model. Other studies also found no association [7, 25, 28, 29].

#### 3.3.9. Corticosteroids use

Two studies have discussed the effect of corticosteroids on the risk of COVID-19 and none found any association between the infection and using corticosteroids in the past 3 months [24], pulse therapy [18], or treatment with prednisolone [18].

Three studies have discussed the association of methylprednisolone use and the outcome of COVD-19. The results of COViMS registry showed an increased risk of hospitalization [OR 2.62 (1.33-5.17)] and death [OR 4.17 (1.13-15.4)] in patients who received glucocorticoid in the past 2 months [7]. MUSC-19 registry also found an association between methylprednisolone use in the past 1 month and worse outcome of the infection [OR 5.24 (2.20–12.53)] [6]. Another study assessing methylprednisolone use in the past 1 month found no association [29].

#### 3.3.10. COVID-19 recommendations

Three of four studies supported that contact with a confirmed COVID-9 case is a risk factor of COVID-19 susceptibility [OR 4.38 (1.04, 18.54) [19], OR 27.29 (6.28, 118.66) [17], OR=197.02 (56.36, 688.79) [24]]. Of them, two studies [17, 24] found it as an independent risk factor, but another just applied a univariate model [19]. Two studies provided no support for an association between COVID-19 and history of contact with a person with respiratory symptoms [17, 24]. In contrast, a single-center study suggested that a history of contact with a person with respiratory symptoms is an independent risk factor of COVID-19 [OR 7.23 (3.48, 15.02)], but not reporting recent contact with a patient with COVID-19 diagnosis [22].

There was no association between the risk of being infected with COVID-19 and observing quarantine recommendations [22] or adhering to social distancing recommendations [19]. however, one study suggested that self-isolation patients were less likely to be suspected for COVID-19 [OR 0.064 (0.016 to 0.259)], but not be confirmed [15].

#### 3.3.11. Socioeconomic status

Two studies have discussed socioeconomic status, one multi-centric study used Area Deprivation Index (ADI) and years of education [19], and another study from Spain used annual net wage [24]. None found any relation between indicators of SES and COVID-19 susceptibility. No study has investigated the effect of SES on the severity of COVD-19.

#### 3.3.12. Vitamin D levels

No study examined the association of vitamin D levels with the risk of contracting COVID-19. Two studies found no association between vitamin D statuses and the outcome of COVID-19 found [17, 24].

#### 3.3.13. Other factors

One study suggested that patients with depression were more likely to developed COVID-19 [16]. There was an association between COVID-19 and the duration of current DMT, but it didn’t remain significant in the multivariate model [24]. No association between COVID-19 and lymphopenia [24], IgG hypogammaglobulinemia [24], working on site [19], number of times leaving home [19], number of times household contacts leaving home [19], viral symptoms [19], relapse in previous two weeks [16], urgent care or emergency room visit [16], hospitalization [16], Charlson comorbidity index [16], working as health professionals [14] were observed. There was also no association between severity of COVID-19 and current DMT duration [24] and lymphopenia grades [24].

## Discussion

In this systematic review, we aimed to summarize the existing evidence on risk factors for COVID-19 susceptibility and severity in patients with MS. The findings of this study support the theory that MS patient’s demographic characteristics, clinical features, DMTs exposure, and observing COVID-19 recommendation could be linked with risk of susceptibility for COVID-19 and severity of the infection.

Our study suggests that aging could be an independent risk factor for poor COVID-19 outcomes in MS patients. This is in accordance with the results of COVID-19 studies in the general population suggesting a role of aging in the severity of COVID-19 [31-33]. It is estimated that each five-year increase of age in patients aged≥ 10 years is associated with a mean increase in infection fatality rate (IFR) of 0.59% [34]. On the other hand, some studies have found that MS patients with younger age are more susceptible to COVID-19 [15, 24]. It has been also suggested a reduced risk of COVID-19 susceptibility in patients with the progressive course [14, 15, 23]. This could be because behavioral and clinical factors could decrease the odds of COVID-19. Older patients are more disable and less involved in high-risk activities such as smoking or alcohol consumption, going to travel, working outside the home, and spending a long time in social interaction. As a result, they may stay at home and not be in close contact with COVID-19 cases, which would reduce cases among older and disable MS patients.

It is generally believed that men are more likely to be infected with SRAS-COV2 and develop severe COVID-19 illness [35, 36]. However, no strong evidence regarding the independent association of sex with COVID-19 susceptibility among MS patients was found. On the other hand, it seems that the male gender may have an association with more severe COVID-19 and mortality [5, 7, 30]. Men patients are more likely to have aggressive MS and worse clinical disability [37], which can increase the risk of infection compared to women [38]. It is a need for sex-stratified analysis to determine the possible effect of sex on COVID-19 outcomes and mortality.

The existing evidence suggests that MS patients with greater disease severity may be at higher risk of poor COVID-19 outcomes [5, 7, 27, 29]. However, it seems that the severity of MS is not associated with COVID-19 susceptibility. More advanced disease-related functional limitations such as respiratory muscle weakness, impaired cough, and respiratory failure [39-41] can increase the risk of respiratory infections such as COVID-19 [42, 43]. It seems that disease duration and course of the disease are not independent risk factors for COVID-19 severity. The association of course and duration of MS could be related to age, MS severity, and presence of comorbidity.

There is much less information about the effects of race and ethnicity on COVID-19 susceptibility and severity among MS patients. Previous studies have found disproportionality of COVID-19 cases among different races and ethnicity; however, the difference was not statistically significant [15, 16, 19, 24]. In the report from the North American registry, the Black or African American race was independently associated with adverse COVID-19 outcomes [7]. It has been proposed that the impact of race and ethnicity is relating to preexisting comorbidity, healthcare access, behavioral, and socioeconomic factors rather than genetic background [44-46].

Our findings suggest that obesity could be a driver of COVID-19 poor outcomes [5, 7, 27]. The results of COViMS and Covisep registries provided some evidence regarding the association of cardiovascular comorbid conditions and adverse disease prognosis [5, 7]. Moreover, COViMS results suggested that hypertension may be linked to an increased risk of death [7]. The possible mechanism underlying the associations is expression of angiotensin-converting enzyme 2 (ACE2) in myocardial cells and adipocytes of obese, which facilitates entry of SARS-COV2 into heart cells and adipocytes [47-49]. Further studies are required to assess the relation of other specific comorbidities with COVID-19 susceptibility and severity among MS patients.

Association between corticosteroids and COVID-19 susceptibility among MS patients have been investigated in two studies but none was able to find any relation [18, 24]. However, two large studies provided evidence suggesting that previous use of corticosteroids could affect COVID-19 prognosis [6, 7]. This is in accordance with previous studies that showed patients with rheumatoid arthritis treating with glucocorticoid were at higher risk of worse COVID-19 outcomes [50-52]. A possible explanation for these results may be the immunosuppressive effects of glucocorticoids which predispose the patients to various infections and worse outcomes [53]. The important point that must be taken into consideration is that administration of corticosteroids in patients with severe or critical COVID-19 could be associated with better outcomes [54, 55]. These suggest that the timing of corticosteroid exposure is a crucial issue that must be attended.

Some results of the published studies concluded that CD20 B-cell depleting drugs including rituximab and ocrelizumab may predispose MS patients to COVID-19 [21-23] and make them more vulnerable to develop severe infection [6, 7]. A recent systematic review investigating COVID-19 characteristic among MS patients showed that patients received anti-CD20 agents had highest hospitalization and mortality rates among DMTs [9]. This goes in line with studies on inflammatory rheumatic and musculoskeletal diseases which found an association between rituximab and critical COVID-19 outcome [52, 56, 57]. Although the exact reason for this association is elusive, it is suggested that patients treated with anti-CD20 monoclonal antibodies have decreased antibody production which can lead to an impaired immune response to SARS-COV2 [58-60].

As far as current evidence shows, it can be suggested that other DMTs probably are associated with neither increased nor decreased risk of COVID-19 susceptibility and disease severity. However, there is some evidence regarding the beneficial effect of IFN [16] and GA [21] on the risk of being suspected for COVID-19. As we were writing this article, Sormani et al. reported an update on the results of the MUSC-19 study suggesting a decreased risk of severe COVID-19 among patients treating with interferon compared to those on DMF as a reference group [61]. The beneficial effect of IFN and GA on COVID-19, in general, remains unclear [62]. Therefore, we are unable to draw a certain conclusion regarding the association of IFN and GA with COVID-19.

The highest hospitalization and mortality rates of COVID-19 were reported among patients who received no DMT [9]. Our study suggested that receiving no DMT may be associated with COVID-19 severity, but probably not as an independent risk factor [5, 6, 24]. Patients with older age and advanced terminal stage, where the risks of treatment overweight the benefits, most don’t receive DMT. It can be therefore suggested that older age, greater disease severity, and more frequent comorbidities put these patients at increased risk of developing severe COVID-19.

Our review suggests that a history of contact with a COVID-19 case is associated with an increased risk of being suspected for COVID-19 [17, 19, 24]. This result is reasonable because SARS-COV2 transmission mainly occurs during close contact with an infected case by airborne mechanisms or saliva and respiratory secretions [63]. Such a result emphasizes the importance of observing COVID-19 recommendations to reduce the risk of acquiring COVID-19.

Our study has some limitations. Due to considerable differences in the definition of susceptibility for COVID-19, the severity of the infection, and analysis methods, we were unable to quantitatively synthesize the results. We were unable to assess the strength of association. We included conference abstracts that have not been reviewed properly and the validity of the results is questionable. However, the inclusion of abstracts can be associated with decreased risk of publication bias. Another limitation of this study is exclusion of non-English studies. Several other risk factors likely exist, however, we only included factors reported in studies on MS patients. Some factors were interpreted as not being independent risk factors for COVID-19 susceptibility or severity. It is worth noting that this systematic review is based on the current published articles and conference abstracts, some of which were relatively small or didn’t have the necessary statistical power. Therefore, caution must be used when interpreting the association of each factor with COVID-19 susceptibility or severity.

In conclusion, factors associated with COVID-19 susceptibility and outcomes are accordance with those reported from the population of general COVID-19 patients. Moreover, DMTs including interferons, glatiramer acetate, and CD20-depleting agents could be associated with COVID-19. Our knowledge of factors associated with COVID-19 susceptibility and severity among MS patients needs to progress to design appropriate programs from preventing infections of MS patients to identify high-risk patients early.

## Data Availability

Data available on request from the authors

